# SARS-CoV-2-antibody response in health care workers after vaccination or natural infection in a longitudinal observational study

**DOI:** 10.1101/2021.06.09.21258648

**Authors:** Jonas Herzberg, Tanja Vollmer, Bastian Fischer, Heiko Becher, Ann-Kristin Becker, Human Honarpisheh, Salman Yousuf Guraya, Tim Strate, Cornelius Knabbe

## Abstract

**Background:** Following a year of development, several vaccines have been approved to contain the global COVID-19 pandemic. Real world comparative data on immune response following vaccination or natural infection are rare.

**Methods:** We conducted a longitudinal observational study in employees at a secondary care hospital affected by the COVID-19 pandemic. Comparisons were made about the presence of anti-SARS-CoV-2 immunglobulin G (IgG) antibody ratio after natural infection, or vaccination with one or two doses of BioNTech/Pfizer (BNT162b2), or one dose of AstraZenca (Vaxzevria) vaccine.

**Results:** We found a 100% humoral response rate in participants after 2 doses of BNT162b2 vaccine. The antibody ratio in participants with one dose BNT162b2 and Vaxzevria did not differ significantly to those with previous PCR-confirmed infection, whereas this was significantly lower in comparison to two doses of BioNTech/Pfizer. We could not identify a correlation with previous comorbidities, obesity or age within this study. Smoking showed a negative effect on the antibody response (p=0.006)

**Conclusion:** Our data provide an overview about humoral immune response after natural SARS-CoV-2 infection or following vaccination, and supports the usage of booster vaccinations, especially in patients after a natural SARS-CoV-2 infection.

## Introduction

Severe acute respiratory syndrome coronavirus type 2 (SARS-CoV-2), which causes the corona-virus-disease-19 (COVID-19) has spread, beginning in 2019 in China, throughout the whole world [1].

Up until May 2021, more than 162 million cases and 3 million deaths related to COVID-19 were reported from around the world [1].

In the global fight against this pandemic, hospitals and health care workers are a core factor [2].

After several lockdowns in almost all affected countries with a persistently high incidence, mass vaccination was seen as one of the most promising elements to control the pandemic. Beginning in December 2020 the first vaccines against COVID-19 were approved worldwide [3–6]. These included the mRNA vaccine from BioNTech/Pfizer (BNT162b2) and the vector-based vaccine by AstraZenca (Vaxzevria), which were the first two vaccines approved in Germany.

Both vaccines use a two-step approach with a primary and then booster dose after several weeks [4]. With regards to BioNTech/Pfizer, the time period between the two doses is recommended as 21 - 42 days, whereas it should be between 8 - 12 weeks with AstraZeneca.

Due to limited availability of vaccines, the German Standing Committee on Vaccination (STIKO) recommended a prioritized procedure [7]. Because of their important role within the pandemic and their higher risk for occupational exposure, hospital employees were grouped in the highest prioritization group for vaccination, and vaccination began in the end of December 2020.

Initial data showed a high effectiveness of the vaccines in preventing not only the symptoms but also the transmission of the SARS-CoV-2 virus even after the first dose [3,8–12]. Both vaccines act by inducing an immune response against the S1 spike protein [13]. This antibody response is shown to correlate with the functional viral neutralization [14,15].

Until now, real-world data about the humoral immune response and potential factors causing a reduced antibody reaction are rare, especially for highly affected and important groups such as health care workers [14,16].

After starting our seroprevalence study in April 2020 at Krankenhaus Reinbek St. Adolf-Stift, a German secondary care hospital, we collected additional blood specimens one year later from study participants. This cohort included participants after natural SARS-CoV-2-infection, one or two doses of vaccination or those reported to be infection-and vaccination-naive. Therefore, semiquantitative IgG antibody ratios against SARS-CoV-2 spike S1 protein were categorized as being after a single or two doses of BioNTech/Pfizer vaccine, one dose of AstraZeneca, post-infectious participants and participants without vaccination or reported infection. All participants reported their symptoms or side effects in a questionnaire. The aim of this study, of a well-defined cohort after different immune stimulations, was to investigate the anti-SARS-CoV-2 antibody ratio and asses further correlation with possible co-factors such as age, gender or previous medical history.

## Methods

### Study design

The “Prospective Sero-epidemiological Evaluation of SARS-CoV-2 among Health Care Workers” (ProCoV-study) was a longitudinal trial started in April 2020 [17]. The study center is a secondary care hospital located in the province of Schleswig-Holstein near the border of the city of Hamburg in Northern Germany. It functioned as a core facility during the pandemic, treating more than 250 PCR-confirmed COVID-19-patients on isolation wards and in the intensive care unit.

All hospital employees were invited to participate during the first phase of the pandemic, so that the longitudinal trial began in April 2020. During that phase, a longitudinal evaluation of seroprevalence and PCR-positivity was performed followed by a half-year seroprevalence evaluation in October 2020 [18].

In December 2020, after the first COVID-19-vaccinces were approved, the healthcare workers were offered vaccination using either the mRNA vaccine of BioNTech/Pfizer or the vector-vaccine from AstraZeneca. In total 709/1050 (67.5%) employees received at least one dose of vaccine before this study and 133/1050 (12.7%) tested positive for SARS-CoV-

2. The majority of participants vaccinated by BioNTech/Pfizer got their booster dose within 42 days, whereas the booster dose of AstraZeneca was not given prior to the blood testing for this study. Participants with a documented COVID-19 infection within the last 6 months were excluded from vaccination, as advised by the Robert-Koch-Institute [7].

All blood samples were collected between April 19^th^ - 30^th^ 2021 and stored at 4°C.

The antibody-testing was fully automated performed using the semiquantitative anti-SARS-CoV-2-ELISA (IgG) from Euroimmun (Lübeck, Germany) detecting the S1 domain of the SARS-CoV-2 spike-protein with, according to the manufacturer, a specificity of 99.0% and sensitivity of 93.8% [19]. This test was used during all phases of this trial to ensure longitudinal comparability. In accordance with the manufactural advice, a ratio below 0.8 was considered negative, a ratio ≥0.8 to <1.1 was considered equivocal, and a ratio ≥1.1 was considered positive.

In addition to the initial assessment in April 2020, a questionnaire regarding symptoms of a previous SARS-CoV-2 infection, post-vaccination symptoms, and reasons for refusal of a vaccination (if applicable) was collected during this time period. By describing their individual symptoms, the severity of the SARS-CoV-2-infection could be retrospectively graded by the participants into mild, moderate or severe based on their personal rating.

All study activities were conducted in accordance with the Declaration of Helsinki. Written and informed consent was given by all study participants prior to enrolment, and the Ethics Committee of the Medical Association Schleswig-Holstein approved this study. It was prospectively registered at the German Clinical Trial Register (DRKS00021270).

### Statistical analysis

IBM SPSS Statistics Version 25 (IBM Co., Armonk, NY, USA) was used for statistical analysis.

All variables are presented as means or medians with standard deviation. Categorical variables are shown as numbers with percentages. Fisher’s exact test or chi-square test was used to determine relationships between categorical variables depending on size of groups. Exact 95% confidence intervals were provided where appropriate. Differences between groups were analyzed using Man-Whitney-U-test or Kruskal-Wallis-test. A linear regression analysis was done to investigate the joint effect of age, vaccination or previous infection, sex and current smoking on antibody response. A p-value < 0.05 was considered statistically significant.

## Results

In total, 562 of 871 (64.5%) participants provided blood specimens and a completed questionnaire for this follow-up. Among the participants in this follow-up were 434 (77.2%) females and 128 (22.8%) males. The mean age was 43.5 years (± 13.77 years). The main characteristics (mean age, sex, comorbidities, BMI, smoking) remained unchanged to the initial phase of this trial [17].

In the initial assessment, 318/562 (55.5%) participants reported no previous significant medical history.

Characteristics of the study cohort are shown in **Table 1** grouped in accordance to their history of type of immunization.

**Table 1:** Characteristics of the participants in this study M: mean; SD: standard deviation

|  | Total | BioNTech |  | AstraZeneca | Post-infection,<br>no vaccination | No infection or<br>vaccination |
| --- | --- | --- | --- | --- | --- | --- |
|  |  | 1. Dose | 1. and 2. Dose | 1. Dose |  |  |
| N | 562 | 9 | 315 | 117 | 52 | 69 |
| Age (years) M ± SD | 43.5 ± 13.77 | 43.00 ± 11.89 | 44.60 ± 13.65 | 42.56 ± 12.91 | 39.71 ± 12.73 | 43.33 ± 16.21 |
| Sex |  |  |  |  |  |  |
| Male, n (%) | 128 (22.8) | 3 (33.3) | 81 (25.7) | 27 (23.1) | 13 (25.0) | 4 (5.8) |
| Female, n (%) | 434 (77.2) | 6 (66.7) | 234 (74.3) | 90 (76.9) | 39 (75.0) | 65 (94.2) |
| SARS-CoV-2-Infection, n (%) | 65 (11.6) | 4 (44.4) | 6 (1.9) | 3 (2.6) | 52 (100) | 0 (0) |
| Medical history |  |  |  |  |  |  |
| Cardiac, n (%) | 93 (16.6) | 0 (0) | 63 (20.0) | 16 (13.7) | 2 (3.8) | 12 (17.4) |
| Pulmonary, n (%) | 57 (10.2) | 1 (11.1) | 31 (9.8) | 18 (15.4) | 2 (3.8) | 5 (7.2) |
| Metabolic, n (%) | 73 (13.0) | 1 (11.1) | 43 (13.7) | 14 (12.0) | 4 (7.7) | 11 (15.9) |
| Other, n (%) | 103 (18.4) | 1 (11.1) | 60 (19.0) | 18 (15.4) | 6 (11.5) | 18 (26.1) |
| Immunosuppression, n (%) | 14 (2.5) | 0 (0) | 7 (2.2) | 3 (2.6) | 0 (0) | 4 (5.8) |
| Smoking, n (%) | 145 (26.0) | 1 (11.1) | 79 (25.1) | 32 (27.4) | 10 (19.2) | 23 (33.3) |
| BMI, M ± SD | 25.86 ± 6.01 | 23.39 ± 9.55 | 26.03 ± 5.47 | 25.65 ± 7.25 | 25.26 ± 6.22 | 26.23 ± 5.45 |
| IgG test result |  |  |  |  |  |  |
| Positive, n (%) | 466 | 8 (88.9) | 315 (100.0) | 101 (86.3) | 41 (78.8) | 1 (1.4) |
| Equivocal, n (%) | 12 | 0 (0) | 0 | 5 (4.3) | 6 (11.5) | 1 (1.4) |
| Negative, n (%) | 466 | 1 (11.1) | 0 | 11 (9.4) | 5 (9.6) | 67 (97.1) |
| SARS-CoV-2-IgG ratio, M ± SD | 5.62 ± 3.32 | 2.44 ± 1.23 | 8.08 ± 1.04 | 3.65 ± 2.42 | 2.83 ± 2.33 | 0.19 ± 0.19 |

65 participants (11.6%) reported a previously PCR-confirmed SARS-CoV-2 infection between April 2020 and April 2021, of which 60 (92 %) occurred in the last six months of the period.

324 participants received at least one dose of BioNTech/Pfizer vaccine and 117 participants one dose of AstraZeneca. 52 participants reported a former SARS-CoV-2 infection followed by no vaccination, and 69 participants were not infected and also did not receive a vaccination due to various reasons.

In the group of previously infected participants without subsequent vaccination, the antibody levels correlated significantly with the severity of the symptoms reported (p=0.016) (**Figure 1**).

**Figure 1:**
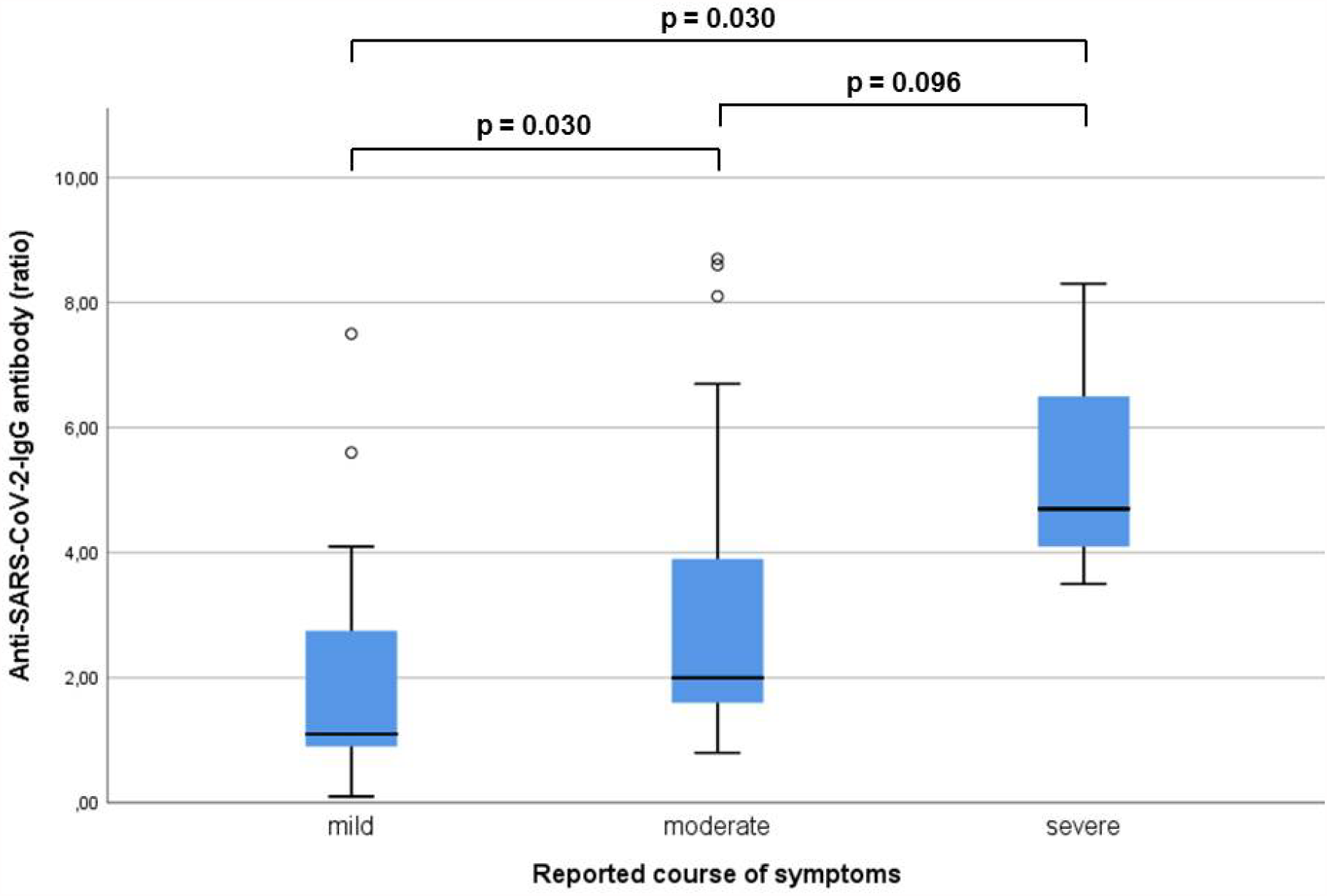
Anti-SARS-CoV-2-IgG antibody ratio in accordance with the subjectively severity of reported symptoms in previously infected participants without vaccination (n=52) Participants with severe symptoms showed a significantly higher anti-SARS-CoV-2-IgG antibody ratio (p=0.030) compared to those with mild symptoms.

The severity of reported post-vaccination symptoms did not correlate with the antibody response after the initial dose of BioNTech/Pfizer (p=0.645) or AstraZeneca (p=0.946), however it could be correlated with the symptoms after the second dose of BioNTech/Pfizer (p=0.006). This could be proven in a linear regression model (R^2^:0.062; p<0.0001; 95%CI 0.128-0.314).

Factors leading to an increased or reduced antibody-response following BioNTech/Pfizer vaccination could not be identified (**Table 2**). In AstraZeneca, female sex was associated with a higher Anti-SARS-CoV-2-IgG antibody ratio than male sex (3.86 ± 2.34 vs. 2.98 ± 2.60, p=0.029). This association did not persist in a linear regression (R^2^:0.23 p=0.099; 95%CI -0.168 – 1.918).

**Table 2:** Correlation of possible factors causing a reduced or increased antibody response. Bold: statistically significant. * Mann-Whitney-U-test Obesity defined as BMI > 30.0 M: mean, SD: standard deviation

|  | **BioNTech/Pfizer** |  | **p-value** |
| --- | --- | --- | --- |
| | N | Antibody ratio, M ± SD | |
| Sex |  |  |  |
| Female | 240 | 8.00 ± 1.28 | 0.154* |
| Male | 84 | 7.70 ± 1.67 | |
| Obesity | 56 | 7.81 ± 1.44 | 0.466* |
| Comorbidity |  |  |  |
| Cardial | 63 | 7.95 ± 1.32 | 0.794* |
| Pulmonal | 32 | 7.95 ± 1.30 | 0.985* |
| Metabolic | 44 | 7.75 ± 1.49 | 0.376* |
| Immunosuppression | 7 | 7.74 ± 1.11 | 0.397* |
| Other | 61 | 7.97 ± 1.34 | 0.759* |
| Smoking | 80 | 7.76 ± 1.48 | 0.114* |
|  | **AstraZeneca** |  | **p-value** |
| Sex |  |  |  |
| Female | 90 | 3.86 ± 2.34 | **0.029\*** |
| Male | 27 | 2.98 ± 2.60 | |
| Obesity | 28 | 3.78 ± 2.47 | 0.792* |
| Comorbidity |  |  |  |
| Cardial | 16 | 3.31 ± 2.32 | 0.229* |
| Pulmonal | 18 | 4.28 ± 2.77 | 0.341* |
| Metabolic | 14 | 3.54 ± 2.66 | 0.727* |
| Immunosuppression | 3 | 3.23 ± 2.76 | 0.776* |
| Other | 18 | 3.39 ± 2.55 | 0.579* |
| Smoking | 32 | 3.10 ± 2.46 | 0.055* |

The highest antibody ratio was measured in participants after 2 doses of BioNTech/Pfizer vaccination, which was significantly higher in comparison to all other groups (p<0.0001). The ratio after a single dose AstraZeneca was significantly higher in comparison to those following natural infection (p=0.014), whereas no significant difference was seen comparing 1 dose AstraZeneca and 1 dose BioNTech/Pfizer (p=0.180) (**Figure 2**). **Figure 3** shows the antibody ratio in accordance to age groups. Only following natural infection, the antibody response differ significantly between these age groups (p=0.030).

**Figure 2:**
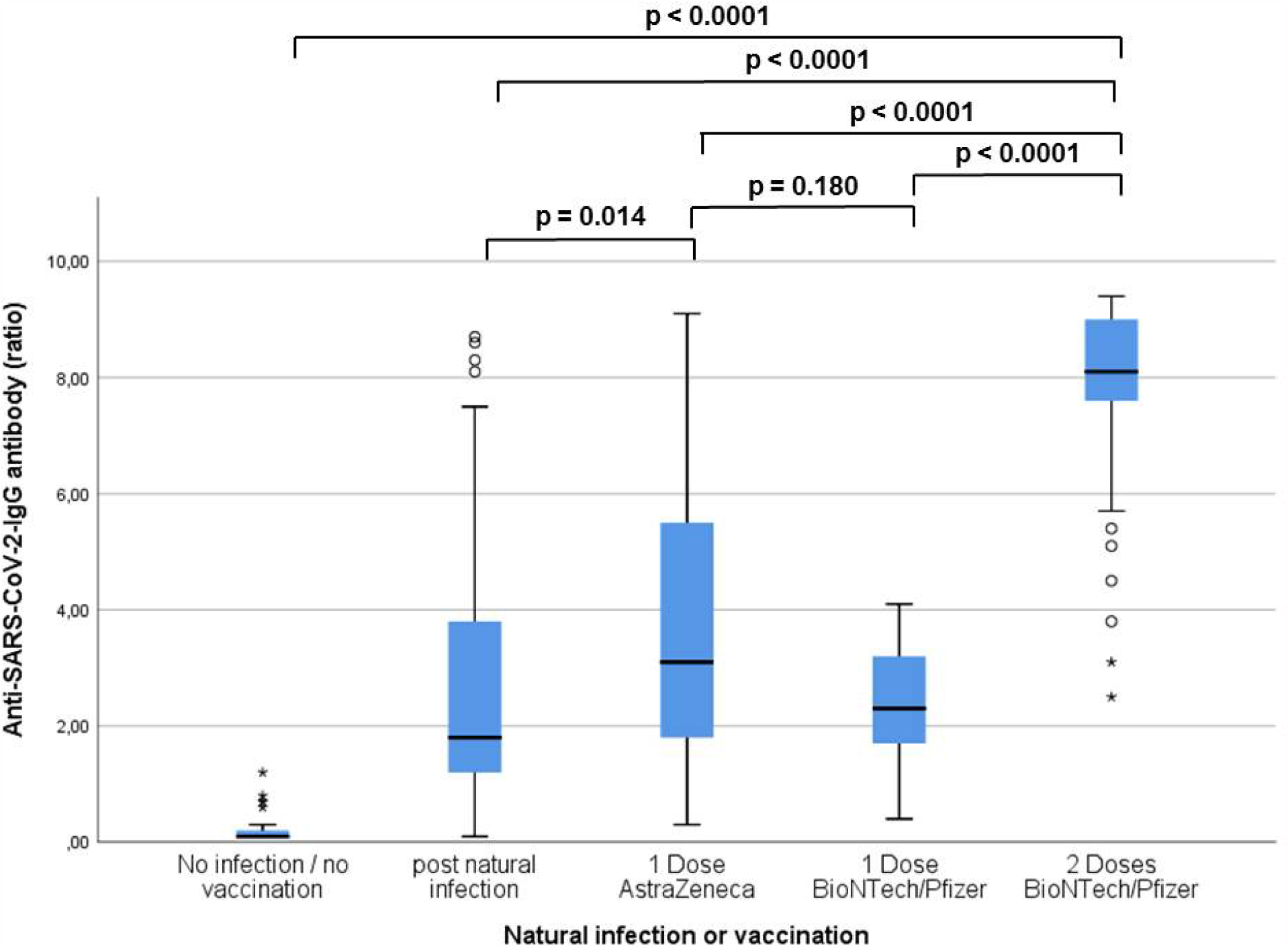
Anti-SARS-CoV-2-IgG antibody ratio according to the type of infection or vaccination in the study group (n=562). Compared to all other groups, participants showed a significant higher antibody ratio after two doses of BioNTech/Pfizer (p<0.0001).

**Figure 3:**
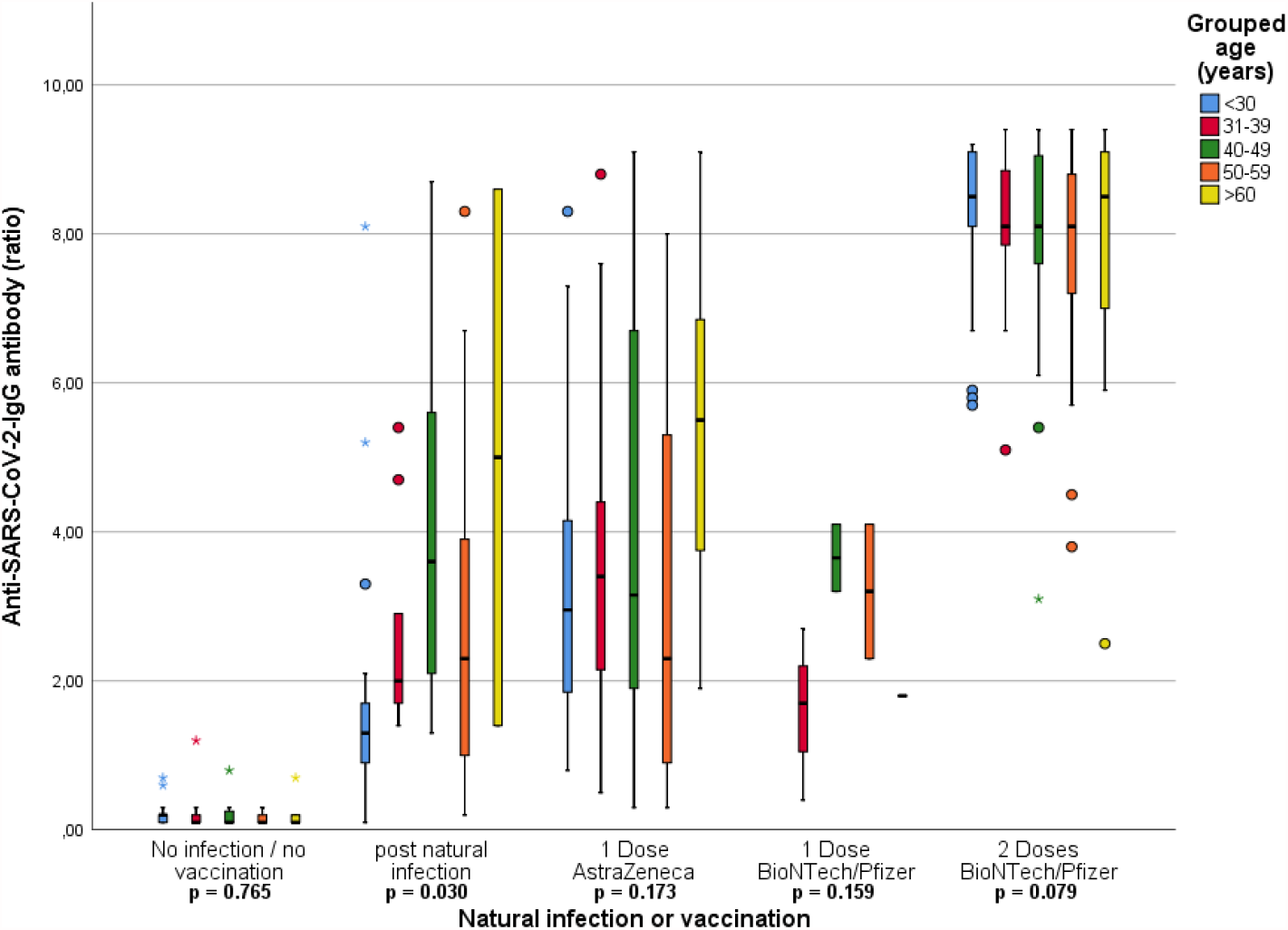
Grouped Age (years) comparing anti-SARS-CoV-2-IgG antibody ratio in relation to natural infection or vaccination (n=562). A significant difference in anti-SARS-CoV-2-IgG antibody ratio could be found for age groups after natural infection (p=0.030), whereas no significant difference could be found within vaccinated groups.

The regression analysis confirmed the univariate analyses regarding the differences in antibody response between the groups. No age effect was observed (p=0.66). The sample size did not allow to investigate differential age effects of antibody response by type of vaccination. A negative effect on antibody response was observed for smokers (p=0.006), and a non-significant negative effect of male sex (p=0.08). Estimates and 95 confidence intervals are given in **Table 3**. Model R^2^ is 0.79.

**Table 3:**
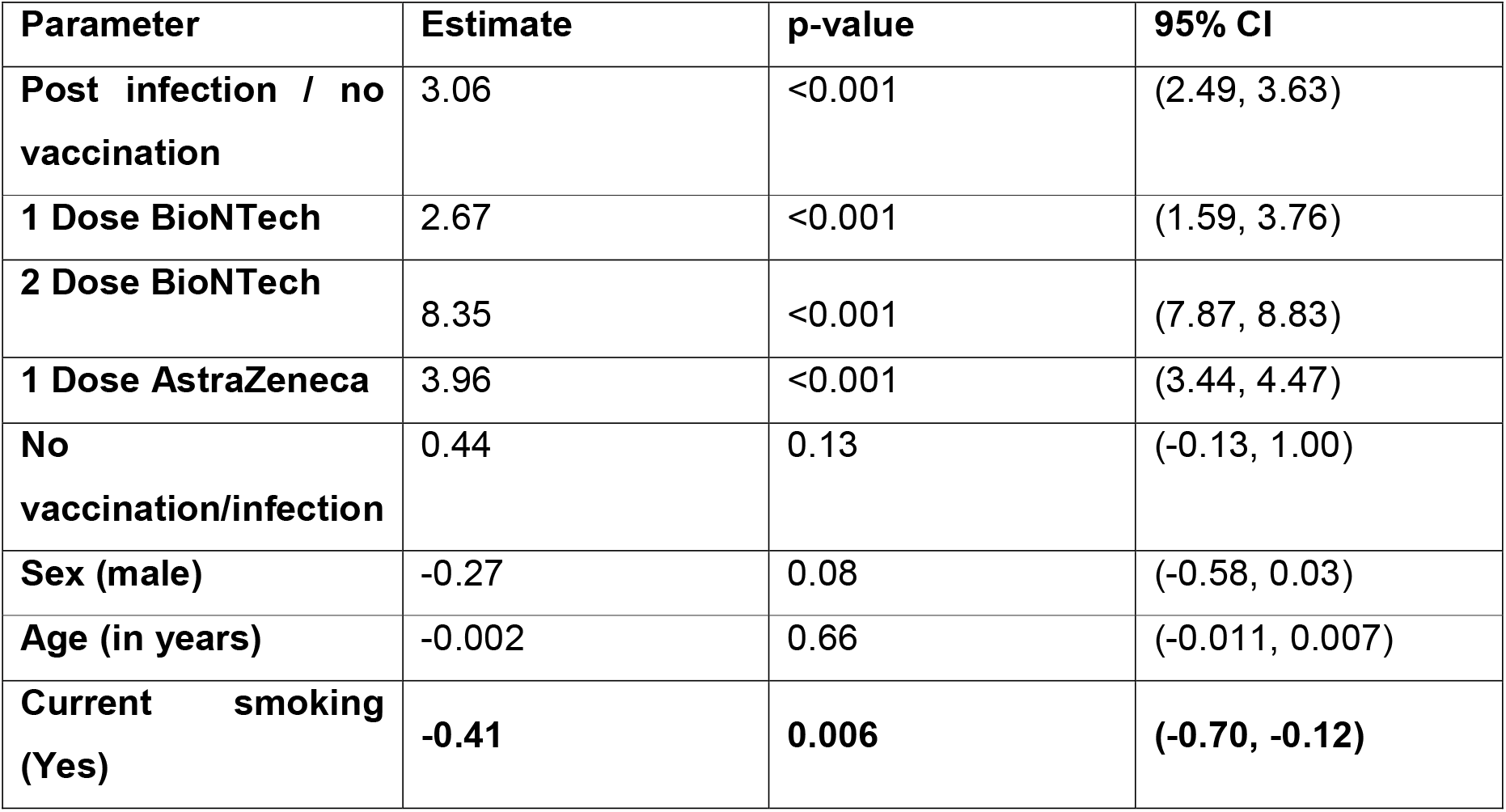
Linear regression for Anti-SARS-CoV-2 antibody ratio as dependent variable and type of immunization and age as covariable CI: confidence interval

## Discussion

This study analyzes the humoral response to natural SARS-CoV-2 infection and different vaccines in a well-defined group of hospital employees. Until now, limited data has been available looking at antibody response to either a single or double dose of BioNTech/Pfizer or AstraZeneca vaccine in comparison to natural infection or immune-naïve people. This study reveals a 100% humoral immune response to two doses of BioNTech/Pfizer vaccine. We found a positive correlation between the anti-SARS-CoV-2 antibody ratio after the second BioNTech/Pfizer vaccination and the number of symptoms reported after this injection. Besides age, no factor causing a reduced immune response could be identified in this trial.

### Factors associated with reduced antibody-response

Several factors have been discussed as possibly influencing immune response after vaccination. Independent of the mechanism of action of the vaccine, individual factors play an important role on the personal response.

Müller et al. showed in their recently published data, a reduced immune response in patients over 80 years in comparison to those below 60 years following BioNTech/Pfizer vaccination against SARS-CoV-2 [20]. This reduced immune response to vaccination in the elderly is well described in the literature [20,21]. Abu Jabal demonstrated in their study a significant decreased antibody response after BioNTech/Pfizer vaccination even in younger people [16]. This age dependency could not be found in our evaluation.

In this evaluation, smoking is the only risk factor for a reduced antibody response following the linear regression analysis.

The effect of smoking is discussed with patients undergoing any kind of vaccination. Studies showed a reduced effectiveness of vaccinations, for example in hepatitis vaccination, due to a general immunosuppression caused by smoking [22,23].

Additionally, studies in patients after organ transplantation have suggested that immunosuppression could led to a reduced antibody response [24]. Without having further details about the severity of the immunosuppression, and given the small number of participants reporting immunosuppression in our cohort, we were unable to validate the previously reported finding of reduced antibody response and immunosuppression.

In addition, obesity reportedly causes a reduced immune response to other vaccinations, such as influenza vaccination, even if the cause for this is not clear yet [25]. The effect of a high body mass index on SARS-CoV-2-vaccination remains unclear. Initial data has shown that a reduced immune response in this group may occur [23]. Even if our data did not support this effect in a larger cohort, obese patients should be under special supervision, as these patients are at high risk for SARS-CoV-2 infection [26].

### Use of booster doses

As also reported by Müller et al., the immune response after a single dose is reduced within all age groups for the BioNTech/Pfizer vaccine [20]. This was also found in our study in the group of AstraZeneca receivers and after a single dose of BioNTech/Pfizer. Nevertheless studies reported a high rate of seroconversion even after a single vaccine-dose against SARS-CoV-2 [27]. The data presented by Parry et al. showed no antibody-response in 13% of individuals after a single dose AstraZeneca in elderly patients. This correlates with our findings, with 9.4% negative and 4.3% equivocal results after single dose of AstraZeneca. Bearing this in mind, the prolonged interval between initial and booster doses of AstraZeneca, as was the norm in countries such as Germany, UK or Israel, might raise the question of whether a single dose serves as sufficient protection for these individuals in a high risk sector such as hospitals [20]. In Antibody response after two doses of AstraZeneca and potentially influencing factors need further evaluation. addition to the time frame of subsequent doses, the use of different vaccines is under current discussion. Currently reported data from Spain showed an increased immune response after combination of AstraZeneca with BioNTech/Pfizer as boost dose [28]. Further details remain unpublished at this time, and more research is needed in this regard.

### Impact of post-vaccination symptoms

There is a broad range of side effects reported after vaccination with BioNTech/Pfizer or AstraZeneca, ranging from local symptoms to systemic post-vaccination symptoms such as fever or headache. These occurred in up to 68.5% of participants after the second dose of BioNTech/Pfizer and up to 58.7% after first dose of AstraZeneca [29]. We found no correlation between the reported severity of post-vaccination symptoms and immune response measured by antibody levels. Müller et al. could not find such a correlation either after the first or second dose of BioNTech/Pfizer vaccine [20]. As shown by Menni et al. the occurrence of reported side effects is more common in women and in younger people [29] which may explain why the majority of participants in our study reported at least one post-vaccination symptom, but these reports are not comparable between individuals.

### Limitation

In this trial, the major limitation is its single-center structure. The study was performed using previously defined timepoints for blood sampling for a longitudinal correlation. Therefore the time frame between vaccination and blood sampling differed between groups of participants. Due to this, one participant after a single dose BioNTech/Pfizer showed no antibody-response, likely because of a short interval between vaccination and blood test (less than 14 days).

Even after a rigorous testing schedule in the study center, asymptomatic infections could be possible. This would provide a possible explanation for one participant without reported SARS-CoV-2 infection or vaccination, with a positive antibody response.

Besides the humoral immune response, cellular response specifically regarding T-cells should be evaluated, especially in participants without an adequate immune response.

Females are highly overrepresented in both groups, representing a common trend in health care workers [30]. Especially in the group of non-vaccinated and non-infected individuals, females are overrepresented. One possibility may be due to the concern of post-vaccination complications regarding fertility.

An additional antibody evaluation after the second dose of the AstraZeneca vaccine, (depending on whether the booster vaccination was using AstraZeneca or BioNTech/Pfizer) could help provide more details on the differences in humoral immune response.

Further evaluations of antibody response after vaccination are needed to investigate the longitudinal persistence of antibodies and the need for further booster vaccinations.

## Conclusion

This study shows a strong immune response in health care workers who received 2 doses of BioNTech/Pfizer vaccination. Participants with a former SARS-CoV-2 infection showed an antibody ratio similar to one dose of BioNTech/Pfizer or AstraZeneca, however, this level depends on the severity of the reported symptoms and time since vaccination.

## Data Availability

The data generated and analysed during this study are available from the corresponding author on reasonable request.

## Acknowledgments

We would like to acknowledge the effort of all team members at the Krankenhaus Reinbek St. Adolf-Stift and at all participating laboratories that analyzed the samples in addition to their daily workload.

Special thanks also to Rebecca Zimmer for her intense linguistic enrichment throughout the entire process of this study.

